# Efficacy of Gene III^®^ L-Ergothioneine Capsules on Postpartum Fatigue, Sleep Quality, and Quality of Life: A Randomized, Double-Blind, Placebo-Controlled Trial

**DOI:** 10.64898/2026.06.08.26355132

**Authors:** Wei Liu, Cong Guo, Wei Ding, Juan Cao, Tong Tong, Shuping Zhou, Feng Shao, Guohua Xiao

## Abstract

**Background:** Postpartum asthenia, characterized by severe fatigue, sleep disturbances, and physiological stress, lacks effective targeted interventions. Ergothioneine (EGT) is a unique, naturally occurring antioxidant that actively accumulates in mitochondria, offering a compelling therapeutic rationale for systemic recovery. This study aimed to evaluate the efficacy of EGT in accelerating postpartum functional restoration and alleviating fatigue.

**Methods:** This single-center, randomized, double-blind, placebo-controlled trial enrolled 40 postpartum women (SF-36 total score ≤ 70) who had ceased breastfeeding. Participants were randomized (1:1) to receive either 120 mg/day of EGT or a matched placebo for 30 days. Efficacy was assessed using the SF-36, Pittsburgh Sleep Quality Index (PSQI), Fatigue Scale-14 (FS-14), and Traditional Chinese Medicine (TCM) asthenia scale. To rigorously evaluate the treatment effects, advanced statistical modeling, including Linear Mixed-Effects Models (LMM) and Analysis of Covariance (ANCOVA) adjusted for baseline covariates, was employed.

**Results:** All 40 participants completed the trial. The EGT group demonstrated robust and accelerated functional recovery. Notably, significant improvements in sleep quality (*p* = 0.0361) and systemic fatigue (*p* = 0.0059) were observed as early as Day 15. Importantly, EGT yielded a statistically significant between-group superiority in alleviating mental fatigue compared to placebo at Day 15 (*p* = 0.0313). By Day 30, the EGT cohort exhibited substantial within-group improvements across all primary metrics, including SF-36 (+35.94%, *p* = 0.0006) and FS-14 (−27.78%, *p* = 0.0011). Furthermore, as an additional physiological benefit, EGT induced a selective and significant reduction in hepatic transaminases (ALT: −30.42%; AST: −17.44%) within normal limits, a trend not observed in the placebo group. EGT was exceptionally well-tolerated with no treatment-related adverse events.

**Conclusions:** EGT supplementation (120 mg/day) safely accelerates postpartum functional recovery, offering rapid relief from mental fatigue and sleep disturbances within 15 days, while concurrently optimizing hepatic physiological status. These preliminary clinical signals warrant confirmation in larger, adequately powered cohorts.

**Trial Registration:** ChiCTR2500114171; Prospectively registered on 2025-12-08.

## 1. Introduction

The postpartum period, often referred to as the ‘fourth trimester,’ represents a critical window of physiological remodeling and psychological vulnerability [1]. A substantial proportion of women suffer from “postpartum asthenia,” a multifaceted syndrome encompassing persistent fatigue, disrupted sleep architecture, and emotional instability [2]. This profound exhaustion stems from a confluence of factors, including severe sleep deprivation, rapid hormonal fluctuations, and sustained neuroendocrine stress [3]. These functional impairments not only compromise maternal quality of life but also potentially interfere with infant caregiving, underscoring the urgent need for effective recovery strategies [4].

Emerging evidence suggests that the systemic stress of childbirth generates a substantial reactive oxygen species (ROS) burden, leading to an acute state of oxidative-redox imbalance [5]. This severe oxidative stress directly impairs mitochondrial energy production, serving as a core biological driver of profound postpartum exhaustion. Central to this transition is the liver, which bears an immense metabolic load as it clears gestational steroid hormones—levels of which drop by over 90 % post-delivery. This heightened oxidative milieu often manifests as subclinical hepatocellular stress, reflected in transient elevations of transaminases such as alanine aminotransferase (ALT) and aspartate aminotransferase (AST) [6]. Given that the depletion of endogenous antioxidants is closely linked to postpartum exhaustion [5], strategies targeting mitochondrial bioenergetics and associated metabolic stress may offer specific therapeutic benefits.

L-Ergothioneine (EGT) is a unique, sulfur-containing amino acid derivative primarily obtained from dietary sources such as mushrooms [7]. Its distinct biological profile relies on a dedicated, high-affinity transporter, OCTN1 (encoded by the SLC22A4 gene), which enables active cellular uptake and tissue-specific enrichment [8]. Unlike conventional antioxidants that rely on passive diffusion, EGT is actively sequestered into mitochondria-rich tissues with high oxidative demands, most notably the liver and the central nervous system, where it serves as a stable and persistent cytoprotectant [9]. Mechanistically, EGT has demonstrated a robust capacity to scavenge hydroxyl radicals and singlet oxygen while preserving mitochondrial membrane integrity [10]. Previous clinical investigations have shown that EGT supplementation at doses ranging from 5 to 25 mg/day can modulate systemic inflammatory markers and attenuate oxidative damage in healthy cohorts [11, 12]. Furthermore, epidemiological data suggest that higher dietary EGT intake is associated with reduced neurodegenerative risk [13] and may promote longevity by upregulating the Nrf2-mediated antioxidant response [14]. These properties make EGT a compelling candidate for mitigating the unique metabolic and oxidative burdens encountered in the postpartum state, potentially translating to an accelerated restoration of physical and mental vitality.

The etiology of postpartum sleep disturbances and chronic fatigue is also intricately linked to the dysregulation of the hypothalamic-pituitary-adrenal (HPA) axis [3]. The abrupt withdrawal of placental hormones, particularly estradiol, significantly impacts hippocampal plasticity and the diurnal cortisol rhythm, contributing to persistent fatigue and mood instability [15]. Normalization of the HPA axis during this period is highly variable and susceptible to oxidative stress and nutritional status [16]. From a Traditional Chinese Medicine (TCM) perspective, these symptoms are often categorized as “Qi-Blood deficiency,” a framework that aligns with modern biological concepts of mitochondrial energy failure (“Qi”) and compromised microvascular nutrient delivery (“Blood”). Integrating validated TCM scales with Western psychometric tools, such as the Fatigue Scale-14 (FS-14), provides a comprehensive, multidimensional evaluation of functional recovery [17].

Despite the strong mechanistic rationale for using EGT to support mitochondrial function and mitigate metabolic stress, its clinical efficacy in the specific context of postpartum recovery remains a significant research gap [11, 12, 18]. To address this, we conducted a randomized, double-blind, placebo-controlled pilot trial to evaluate the impact of a 30-day EGT intervention (120 mg/day) on multifaceted functional recovery. The primary objectives were to assess changes in quality of life (SF-36), sleep quality (PSQI), and fatigue (FS-14), while secondary objectives focused on objective hepatic biomarkers (ALT/AST) as indicators of metabolic safety and physiological stress, and the trajectory of HPA axis normalization. Together, these endpoints aim to provide the first clinical evidence for EGT as a targeted restorative agent for postpartum health.

## 2. Methods

### 2.1 Study Design

This single-center, randomized, double-blind, placebo-controlled pilot trial was conducted at The First Hospital of Anhui University of Science & Technology, China, between December 18, 2025, and February 6, 2026. The study was executed in strict adherence to the Declaration of Helsinki and the International Council for Harmonisation Good Clinical Practice (ICH-GCP) guidelines. The protocol and all associated study materials were thoroughly reviewed and approved by the Institutional Review Board of the participating hospital prior to study initiation. Written informed consent was obtained from all participants before any trial-specific procedures were performed. Reporting follows the Consolidated Standards of Reporting Trials (CONSORT) 2010 guidelines. The trial was prospectively registered at the Chinese Clinical Trial Registry (ChiCTR2500114171).

### 2.2 Participants

Eligible participants were postpartum women who reported clinically significant postpartum asthenia, defined objectively for this study as a baseline 36-Item Short Form Survey (SF-36) total score of ≤ 70. To isolate the direct physiological effects of EGT and eliminate the profound confounding variables of lactation-induced hormonal fluctuations (e.g., prolactin and oxytocin spikes), enrollment was strictly limited to women who had completely ceased breastfeeding. Other exclusion criteria included known hypersensitivity to mushroom-derived products, significant psychiatric diagnoses requiring pharmacological intervention, and clinically unstable hepatic, renal, or cardiovascular diseases.

The sample size of 20 participants per arm was determined based on established recommendations for pilot randomized controlled trials, which typically require 20–30 participants per arm to generate preliminary effect size estimates for future confirmatory trials. Post-hoc power analysis indicated that a future confirmatory trial would require approximately 80–130 participants per arm to achieve 80% power (two-sided α = 0.05) based on the observed effect sizes (Cohen’s d ≈0.30–0.50) for the primary outcome.

A total of 40 participants met the eligibility criteria and were randomized in a 1:1 ratio. The baseline demographic data, including mean age, were statistically comparable between the intervention groups, with a mean age of 31.9 ± 2.43 years for the EGT group and 32.05 ± 3.38 years for the placebo group. Throughout the 30-day intervention period, compliance was exceptional, and no dropouts occurred, resulting in a 100% completion rate (N=40) for the final intention-to-treat (ITT) and per-protocol (PP) analyses (Figure 1).

**Figure 1.**
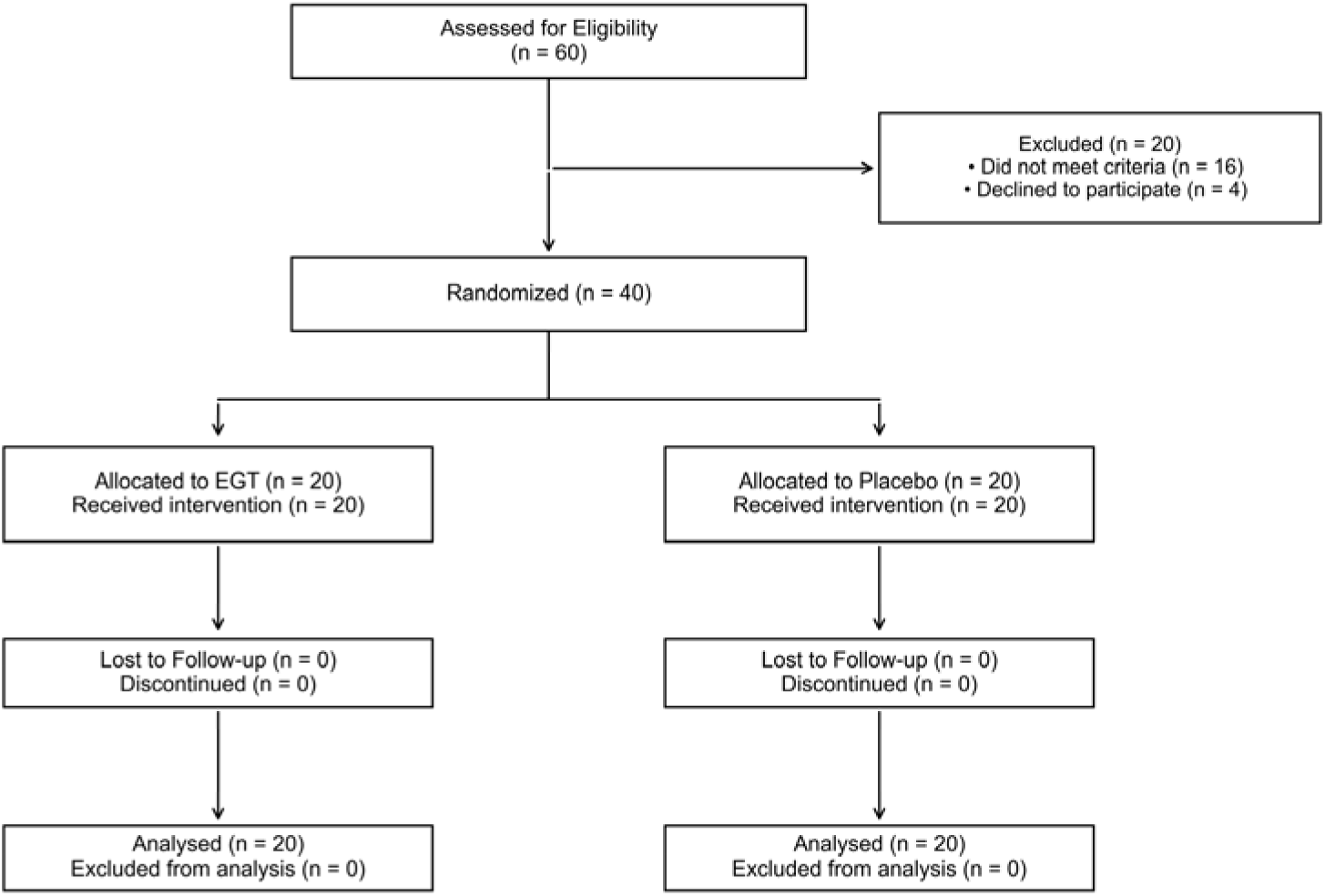
CONSORT flow diagram of the study. This chart illustrates the progress of participants through the recruitment, randomization (1:1), 30-day intervention, and final analysis (n = 40).

### 2.3 Intervention

Participants allocated to the EGT group were instructed to orally administer EGT capsules at a total daily dose of 120 mg (30 mg/capsule, administered as two capsules twice daily). The highly purified Gene III^®^ L-Ergothioneine Capsules (Batch P776678) were manufactured and provided by Jiangsu Gene III Biotechnology Co., Ltd. (Nanjing, China). Participants in the control group received visually and organoleptically identical placebo capsules (Batch 20250301). These placebo capsules were primarily composed of inert microcrystalline cellulose and were specifically manufactured by Xinxiang Lujianyuan Health Technology Co., Ltd. (Xinxiang, China) to match the active intervention.

Given that the primary efficacy endpoints of this trial heavily relied on subjective psychometric evaluations (e.g., SF-36, PSQI), the integrity of the double-blinding procedure was of paramount importance. Therefore, the placebo capsules were meticulously formulated to be indistinguishable from the active EGT capsules, matched precisely not only in size, shape, and weight but also in odor and taste to prevent accidental unblinding. Both the clinical investigators and the participants remained fully blinded to the treatment allocations throughout the 30-day intervention period, and the randomization code was not broken until the completion of the statistical analysis plan.

### 2.4 Outcome Measures

Clinical assessments were systematically performed at three distinct time points: Baseline (Day 0), Mid-point (Day 15), and Study Completion (Day 30). The assessment framework prioritized multidimensional functional recovery as the primary efficacy domain, while treating biochemical markers as indicators of metabolic safety and physiological stress.

#### Primary Efficacy Endpoints

Quality of Life (SF-36): The 36-Item Short Form Survey [19] was employed as the primary metric to assess health-related quality of life. This validated instrument encompasses eight distinct domains: physical functioning, role-physical, bodily pain, general health, vitality, social functioning, role-emotional, and mental health. Scores for each domain and the total score range from 0 to 100, where higher scores reflect superior functional health.

Sleep Quality (PSQI): The Pittsburgh Sleep Quality Index [20] was utilized to evaluate sleep architecture over the preceding month. This 19-item self-rated scale assesses seven components: subjective sleep quality, sleep latency, sleep duration, habitual sleep efficiency, sleep disturbances, use of sleeping medication, and daytime dysfunction. A global PSQI score (0–21) was calculated, with scores > 5 typically indicating significant sleep impairment in clinical cohorts.

Fatigue Assessment (FS-14): Systemic fatigue was quantified using the Fatigue Scale-14 [17]. This instrument distinguishes between physical fatigue (8 items) and mental fatigue (6 items) through a binary scoring system. The total score (0–14) provides a comprehensive measure of exhaustion severity, where higher scores signify more profound fatigue.

TCM Asthenia Scale: A validated Traditional Chinese Medicine (TCM) symptom scale for “Qi-Blood deficiency” was incorporated to capture the specific somatic features of postpartum asthenia. This scale evaluates symptoms such as fatigue, shortness of breath, dizziness, spontaneous sweating, and palpitations (maximum score 57), with score reductions indicating a restoration of the “Qi-Blood” balance.

#### Secondary and Exploratory Endpoints

Hepatic Biomarkers: Fasting serum levels of alanine aminotransferase (ALT; reference range: 7–40 IU/L) and aspartate aminotransferase (AST; reference range: 10–40 IU/L) were measured to monitor objective changes in hepatic metabolic stress and ensure product safety.

Neuroendocrine Marker (Cortisol): To assess the normalization trajectory of the hypothalamic-pituitary-adrenal (HPA) axis, fasting morning blood samples were collected between 07:00 and 10:00. Serum cortisol concentrations (µg/dL) were quantified, with the clinical morning reference range defined as 6.4–22.8 µg/dL.

Depressive Symptoms (EPDS): The Edinburgh Postnatal Depression Scale (EPDS), a 10-item psychometric tool (total score 0–30), was used to screen for emotional distress and depressive symptoms specific to the postpartum period.

### 2.5 Statistical Analysis

Statistical analyses were conducted on an intention-to-treat (ITT) basis. To move beyond simple comparisons and rigorously account for the longitudinal nature of the data, advanced statistical modeling was employed. Data were analyzed using SAS software (version 9.4, SAS Institute Inc.) to ensure regulatory compliance and high-precision computation. Continuous variables are expressed as mean ± standard deviation (SD). The normality of data distribution was rigorously assessed using the Shapiro-Wilk test.

To evaluate the treatment effect over time while accounting for baseline scores and potential confounding, Linear Mixed-Effects Models (LMM) were utilized for all primary clinical outcomes. In these models, treatment group, time (Day 0, 15, and 30), and the treatment-by-time interaction were treated as fixed effects, while participant identity was modeled as a random effect to account for within-subject correlations. This approach allows for a more robust estimation of the acceleration in recovery observed in the EGT group.

Furthermore, Analysis of Covariance (ANCOVA) was performed to compare between-group differences at Day 15 and Day 30. In the ANCOVA models, the respective baseline score, age, and BMI were included as covariates to calculate Estimated Marginal Means (EMMs) and adjusted p-values, thereby isolating the specific effect of EGT supplementation from natural recovery or baseline imbalances.

For secondary physiological indicators (ALT/AST and Cortisol), within-group longitudinal comparisons were performed using paired t-tests or Wilcoxon signed-rank tests. Pearson or Spearman correlation coefficients were calculated to study the coupling between biochemical changes and subjective recovery scores. All statistical tests were two-tailed, and *p* < 0.05 was considered the threshold for statistical significance. Given the exploratory nature of this pilot trial, all reported p-values should be interpreted as nominal signals warranting confirmation in future adequately powered trials.

Additionally, exploratory subgroup analyses were performed to evaluate treatment consistency across baseline demographic strata, specifically focusing on age categories and baseline fatigue severity.

## 3. Results

### 3.1 Baseline Characteristics and Participant Flow

A total of 40 participants were randomized and all 40 successfully completed the 30-day intervention, representing a 100% completion rate for both the intention-to-treat (ITT) and per-protocol (PP) analyses (Figure 1). No participants were lost to follow-up or discontinued due to adverse events.

The baseline demographic and clinical characteristics (Table 1) were well-balanced between the two study arms, with no statistically significant differences across any primary or secondary outcome measures (all *p* > 0.05). Specifically, the mean age was 31.9 ± 2.43 years for the EGT group and 32.0 ± 3.38 years for the placebo group (*p* = 0.87). Body Mass Index (BMI) was also comparable (EGT: 22.34 ± 2.90 kg/m²; Placebo: 21.54 ± 2.42 kg/m²). Baseline physiological biomarkers, including ALT (EGT: 17.75 ± 16.21 IU/L; Placebo: 20.95 ± 19.23 IU/L) and morning serum cortisol (EGT: 11.37 ± 2.85 µg/dL; Placebo: 14.07 ± 3.32 µg/dL), confirmed adequate baseline comparability across key physiological parameters, with the exception of a numerically higher cortisol level in the placebo group. All continuous variables were presented as mean ± standard deviation (SD).

**Table 1.** Baseline demographic and clinical characteristics of the study participants.

| Characteristic | EGT Group (n = 20) | PBO Group (n = 20) | *p*-value |
| --- | --- | --- | --- |
| Demographics |  |  |  |
| Age (years) | $31.9 \pm 2.43$ | $32.0 \pm 3.38$ | 0.87 |
| BMI (kg/m<sup>2</sup>) | $22.34 \pm 2.90$ | $21.54 \pm 2.42$ | 0.35 |
| Baseline Physiological Biomarkers |  |  |  |
| ALT (IU/L) | $17.75 \pm 16.21$ | $20.95 \pm 19.23$ | 0.256 |
| AST (IU/L) | $19.50 \pm 9.02$ | $19.40 \pm 6.62$ | 0.97 |
| Total Bilirubin (µmol/L) | $12.25 \pm 4.10$ | $10.49 \pm 3.90$ | 0.168 |
| Serum Creatinine(µmol/L) | $49.84 \pm 6.47$ | $49.75 \pm 6.98$ | 0.965 |
| Blood Urea Nitrogen (mmol/L) | 4.55 ± 0.95 | 4.72 ± 1.47 | 0.667 |
| Serum Cortisol (µg/dL) | 11.37 ± 2.85 | 14.07 ± 3.32 | 0.08 |

Note: Data are presented as mean ± standard deviation (SD) or n (%). EGT: ergothioneine; PBO: placebo; ALT: alanine aminotransferase; AST: aspartate aminotransferase. P-values reflect baseline comparisons between groups (independent samples t-test for continuous variables and Chi-square test for categorical variables). All differences are statistically non-significant ( $p > 0.05$ ).

### 3.2 Primary Outcome: SF-36 Quality of Life

Both study arms exhibited statistically significant within-group improvements in the SF-36 total score by the end of the 30-day intervention (Figure 2A). In the EGT cohort, the raw total score rose significantly from 46.19 ± 10.98 at Baseline to 62.79 ± 14.45 at Day 30, representing a robust 35.94% relative increase (*p* = 0.0006). The placebo group demonstrated a more modest trajectory (+20.56%, p = 0.0019).

**Figure 2.**
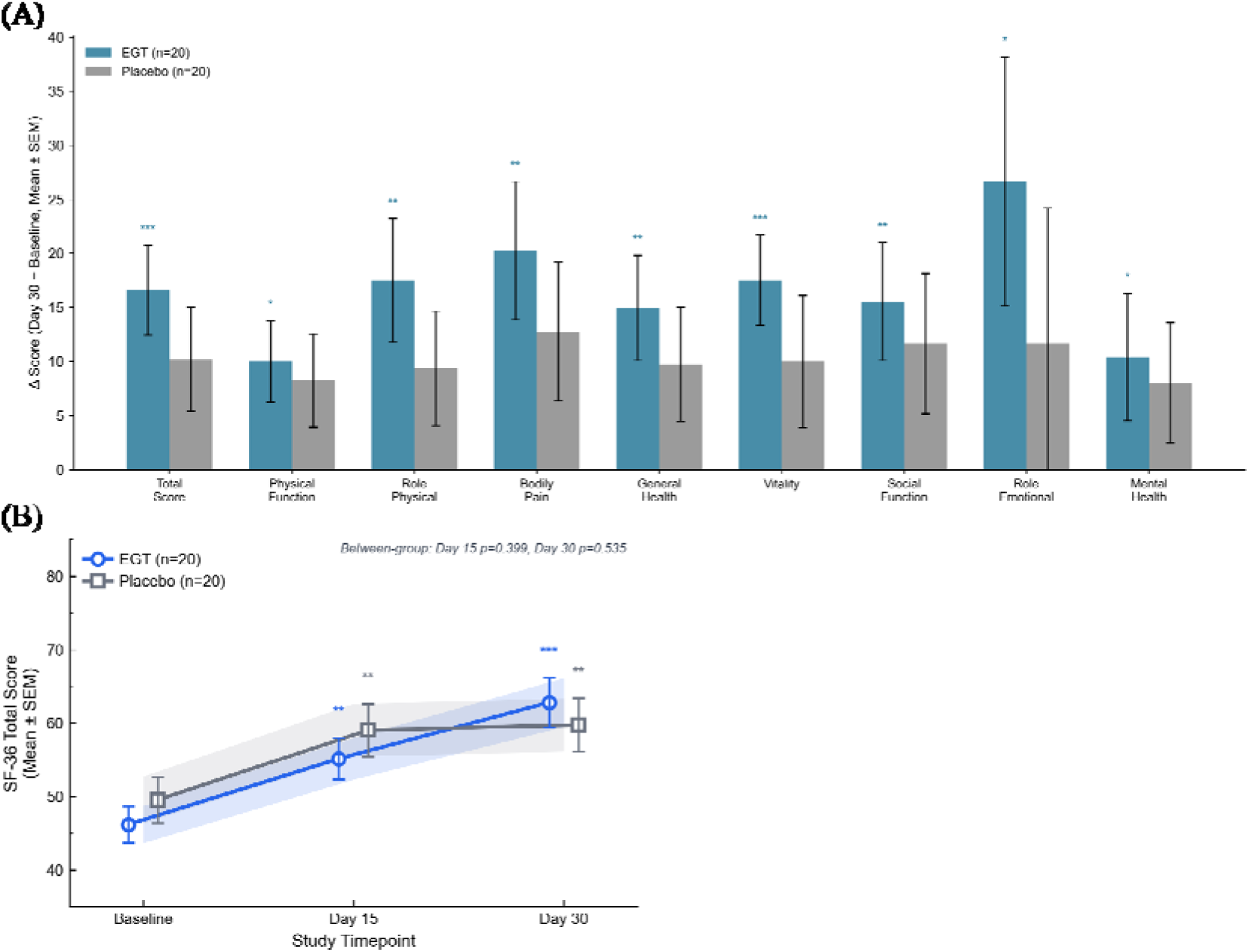
Impact of EGT supplementation on SF-36 Quality of Life. (A) Raw mean SF-36 total scores at baseline and Day 30, showing significant within-group improvements. (B) Longitudinal trajectory of Estimated Marginal Means (EMMs) adjusted for baseline scores, age, and BMI via ANCOVA. Error bars represent standard error (SE). \**p* < 0.05 vs. baseline.

Domain-level analysis revealed that EGT supplementation exerted a comprehensive impact on functional health, with significant within-group improvements achieved across nearly all sub-scales (Table 2). Most notably, the ‘Role-Physical’ (RP) domain showed a striking 147.37% improvement in the EGT group (*p* = 0.0021), far exceeding the natural recovery observed in the placebo group.

**Table 2.**
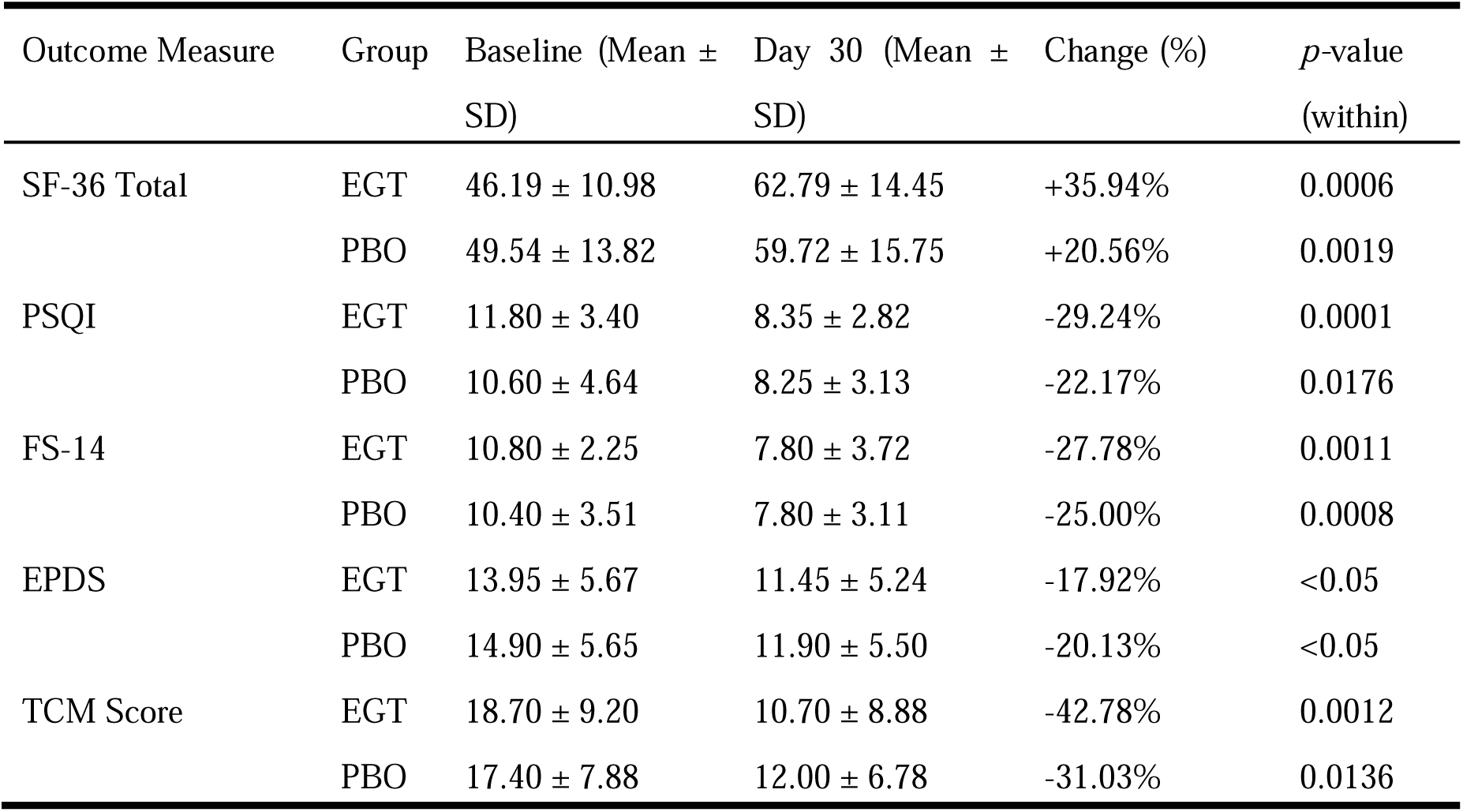
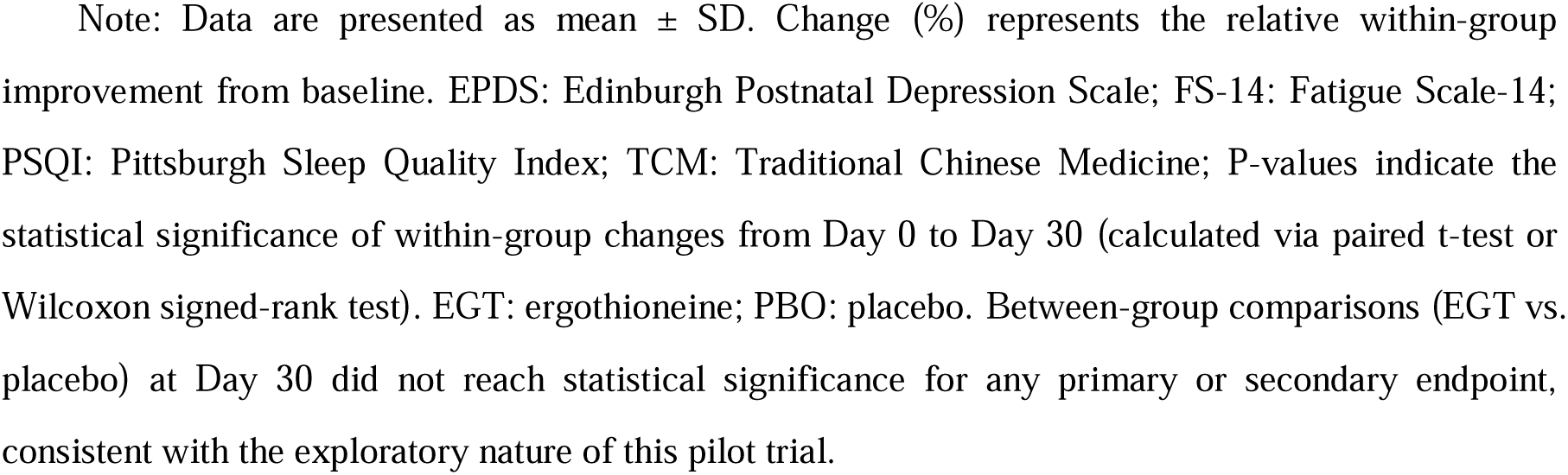
Summary of primary and secondary clinical efficacy outcomes from Baseline to Day 30.

| Outcome Measure | Group | Baseline (Mean ± SD) | Day 30 (Mean ± SD) | Change (%) | $p$-value (within) |
| --- | --- | --- | --- | --- | --- |
| SF-36 Total | EGT | 46.19 ± 10.98 | 62.79 ± 14.45 | +35.94% | 0.0006 |
| | PBO | 49.54 ± 13.82 | 59.72 ± 15.75 | +20.56% | 0.0019 |
| PSQI | EGT | 11.80 ± 3.40 | 8.35 ± 2.82 | -29.24% | 0.0001 |
| | PBO | 10.60 ± 4.64 | 8.25 ± 3.13 | -22.17% | 0.0176 |
| FS-14 | EGT | 10.80 ± 2.25 | 7.80 ± 3.72 | -27.78% | 0.0011 |
| | PBO | 10.40 ± 3.51 | 7.80 ± 3.11 | -25.00% | 0.0008 |
| EPDS | EGT | 13.95 ± 5.67 | 11.45 ± 5.24 | -17.92% | <0.05 |
| | PBO | 14.90 ± 5.65 | 11.90 ± 5.50 | -20.13% | <0.05 |
| TCM Score | EGT | 18.70 ± 9.20 | 10.70 ± 8.88 | -42.78% | 0.0012 |
| | PBO | 17.40 ± 7.88 | 12.00 ± 6.78 | -31.03% | 0.0136 |

Note: Data are presented as mean ± SD. Change (%) represents the relative within-group improvement from baseline. EPDS: Edinburgh Postnatal Depression Scale; FS-14: Fatigue Scale-14; PSQI: Pittsburgh Sleep Quality Index; TCM: Traditional Chinese Medicine; P-values indicate the statistical significance of within-group changes from Day 0 to Day 30 (calculated via paired t-test or Wilcoxon signed-rank test). EGT: ergothioneine; PBO: placebo. Between-group comparisons (EGT vs. placebo) at Day 30 did not reach statistical significance for any primary or secondary endpoint, consistent with the exploratory nature of this pilot trial.

To further evaluate the treatment effect while accounting for longitudinal trajectories and baseline variances, advanced statistical modeling was employed (Figure 2B). Analysis of Covariance (ANCOVA) was utilized to adjust for baseline SF-36 scores (which emerged as a highly significant covariate, *p* = 0.009), age, and BMI. Following this adjustment, the Estimated Marginal Mean (EMM) at Day 30 was 63.48 ± 3.27 for the EGT group compared to 59.03 ± 3.27 for the placebo group. Although this +4.45 point between-group difference did not cross the threshold for statistical significance (Adjusted *p* = 0.3481), the numerical trend consistently favored the EGT intervention throughout the study period. Similarly, Linear Mixed-Effects Modeling (LMM) yielded a treatment-by-time interaction p-value of 0.2038. These findings are consistent with the exploratory design and limited statistical power of a pilot trial (n=40), yet they constitute a positive clinical signal supporting EGT’s efficacy.

Crucially, temporal analysis demonstrated that EGT facilitated an accelerated onset of functional recovery. By the Day 15 mid-point evaluation, the EGT group had already achieved a statistically significant elevation in the SF-36 total score (*p* = 0.0020). In sharp contrast, the placebo group at Day 15 did not exhibit a statistically superior improvement compared to its own baseline. This 15-day acceleration highlights EGT’s unique potential to rapidly shorten the postpartum convalescence window.

Furthermore, exploratory subgroup analyses were conducted to evaluate the consistency of EGT’s therapeutic effects across prespecified baseline characteristics. Consistent with the overall cohort, subgroup analyses revealed that the EGT group demonstrated numerically greater improvements in the SF-36 total score across all evaluated strata, including the 30–35 age demographic (Mean Difference: +10.54, *p* = 0.144). While these between-group differences did not reach statistical significance, the uniform directionality of the effect sizes supports the robustness of the intervention. A detailed visualization of effect sizes across subgroups is provided in Supplementary Figure S1.

### 3.3 Secondary Outcomes: Sleep Quality, Fatigue, Postpartum Depression, and TCM Asthenia

Secondary functional outcomes demonstrated a consistently accelerated recovery pattern unique to the EGT cohort. Regarding sleep architecture, the EGT group’s PSQI global score decreased significantly from a baseline of 11.80 ± 3.40 to 10.50 ± 3.49 as early as Day 15 (*p* = 0.0361), with a further substantial decline to 8.35 ± 2.82 by Day 30, representing a 29.24% relative improvement (*p* = 0.0001). Conversely, the placebo group showed no statistically significant improvement at the Day 15 mid-point (9.95 ± 2.80; *p* > 0.05) and achieved significance only by Day 30 (8.25 ± 3.13; −22.17%, *p* = 0.0176) (Figure 3A).

**Figure 3.**
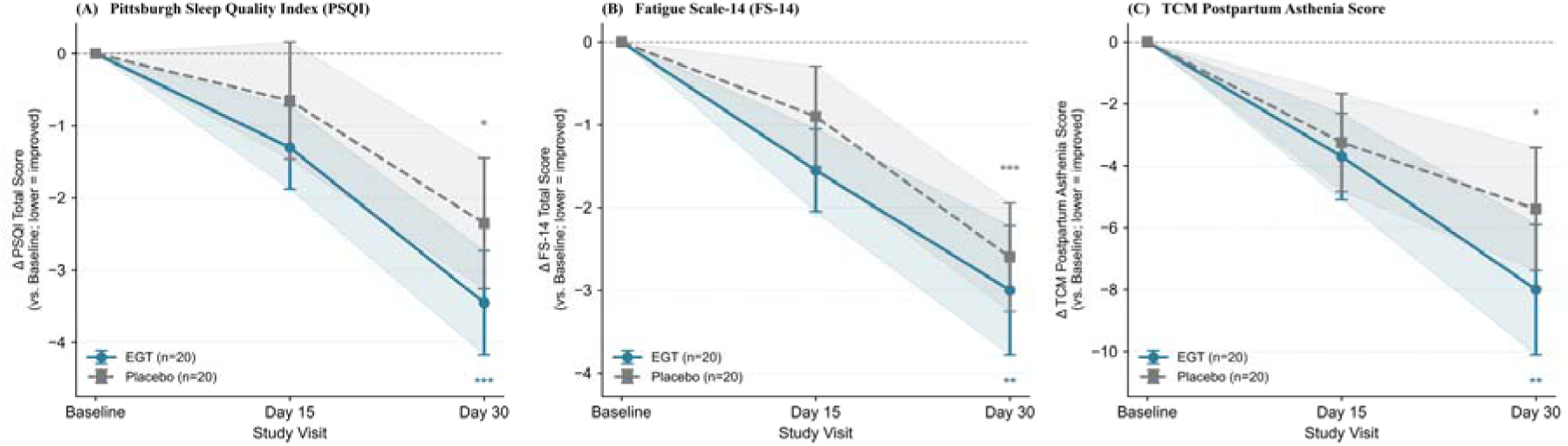
Temporal trajectories of subjective recovery endpoints. (A) PSQI scores, (B) FS-14 scores, and (C) TCM asthenia scores across three timepoints (Baseline, Day 15, and Day 30). Data points represent Mean ± SEM. \**p* < 0.05 indicates a significant within-group change from baseline. Note the earlier onset of significance in the EGT group at Day 15.

Parallel accelerated trajectories were observed for systemic fatigue. EGT supplementation significantly reduced the total FS-14 score from 10.80 ± 2.25 to 9.25 ± 2.68 by Day 15 (*p* = 0.0059), whereas the placebo cohort exhibited no significant change at this early juncture (9.50 ± 3.83; *p* = 0.1543). Crucially, focused sub-scale analysis of the FS-14 revealed a statistically significant between-group difference in the mitigation of mental fatigue at Day 15. The mental fatigue score in the EGT group demonstrated a rapid drop (from 3.80 ± 1.36 to 2.70 ± 1.38), significantly outperforming the placebo group (nominal *p* = 0.0313), which rendered negligible change (−0.05 points). By Day 30, both groups converged at an identical mean FS-14 total score (7.80), strongly suggesting that the distinctive clinical benefit of EGT is primarily characterized by the rapid acceleration of relief from postpartum mental exhaustion in the early phase (Figure 3B).

Postpartum depressive symptoms evaluated via the EPDS showed similar within-group reductions over the 30-day period for both arms (EGT: 13.95 to 11.45; Placebo: 14.90 to 11.90, both *p* < 0.05), with no statistically significant between-group differences detected.

Additionally, the TCM postpartum asthenia score—capturing profound somatic depletion conceptualized as ‘Qi-Blood deficiency’—strongly aligned with Western clinical metrics. In the EGT group, the mean score fell significantly from 18.70 ± 9.20 at baseline to 15.00 ± 8.58 at Day 15 (*p* = 0.0155) and achieved a striking 42.78% reduction by Day 30 (10.70 ± 8.88, p = 0.0012). In contrast, the placebo group demonstrated only a borderline trend at Day 15 (*p* = 0.0538) before reaching significance by Day 30 (−31.03%, *p* = 0.0136). Although formal between-group comparison at Day 30 was not statistically significant, the EGT group exhibited a numerically greater absolute score reduction (8.00 vs. 5.40 points), highlighting its enhanced systemic restorative effects.

### 3.4 Hepatic Biomarkers: Selective Modulation of Transaminases

Among the objectively evaluated physiological biomarkers, EGT supplementation induced a statistically significant and selective within-group reduction in serum transaminases, serving as an additional indicator of metabolic safety and physiological stress mitigation. Baseline ALT and AST values were within normal physiological reference ranges for all participants (ALT: EGT 17.75 ± 16.21 vs. Placebo 20.95 ± 19.23 IU/L; AST: EGT 19.50 ± 9.02 vs Placebo 19.40 ± 6.62 IU/L; both *p* > 0.05), confirming baseline comparability.

ALT levels decreased significantly from 17.75 ± 16.21 IU/L at Baseline to 12.35 ± 6.40 IU/L at Day 30 in the EGT group, representing a substantial 30.42% reduction (*p* = 0.0192). In sharp contrast, the placebo group displayed a non-significant trend in the opposite direction, with ALT levels increasing from 20.95 ± 19.23 to 22.70 ± 30.08 IU/L (+8.35%; *p* = 0.5445) (Figure 4A).

**Figure 4.**
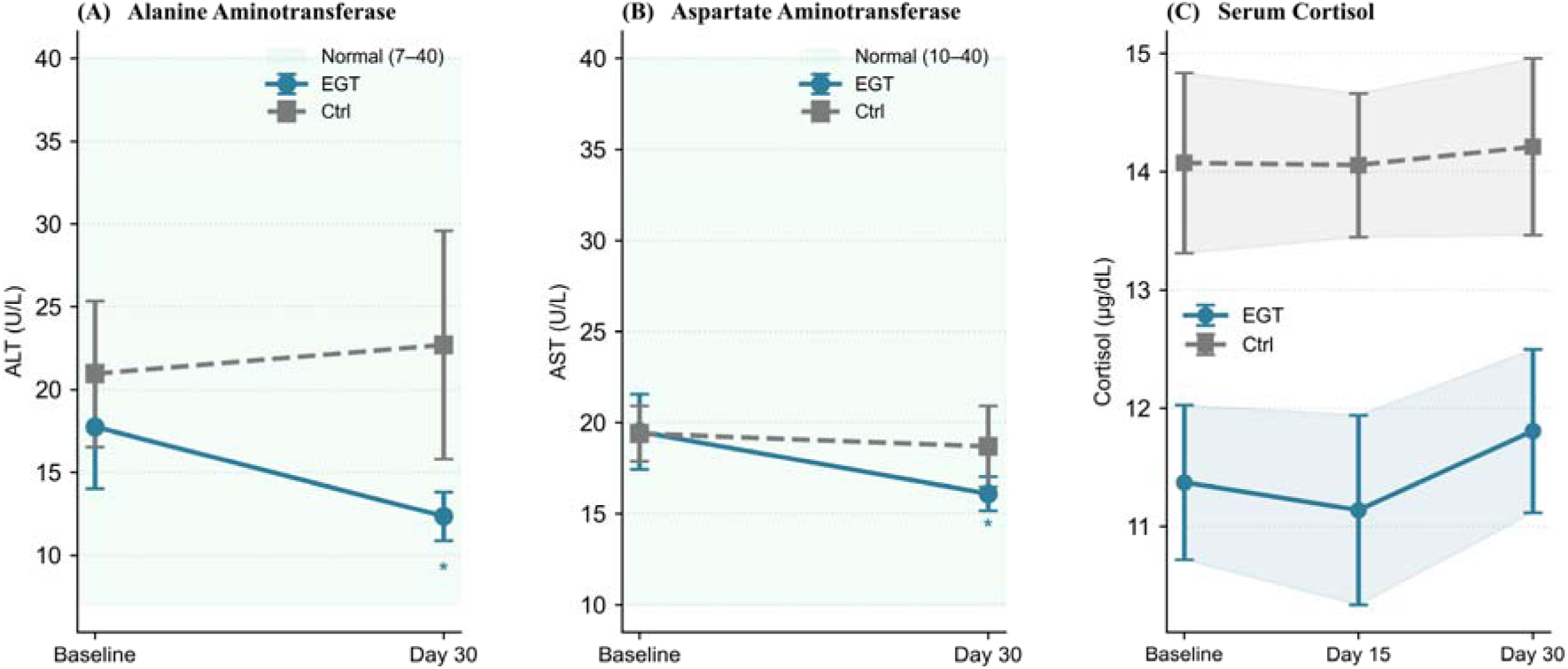
Hepatic transaminase and serum cortisol profiles. (A) Selective reduction of ALT and AST in the EGT group compared to placebo. (B) Stable morning serum cortisol levels within normal reference ranges (6.4–22.8 µg/dL) throughout the study.

Similarly, AST levels decreased significantly from 19.50 ± 9.02 to 16.10 ± 4.09 IU/L in the EGT group (−17.44%; *p* = 0.0058), compared to a clinically negligible reduction in the placebo group (19.40 ± 6.62 to 18.70 ± 9.69 IU/L; −3.61%; *p* = 0.3152). Given the modest sample size and substantial within-group variance—particularly evident in the placebo cohort—this pilot study was inherently underpowered to detect robust between-group differences in metabolic biomarkers at Day 30 (ALT: *p* > 0.05; AST: *p* > 0.05). Nevertheless, the highly divergent physiological trajectories observed—a precise, significant optimization of hepatic transaminases in the EGT arm juxtaposed with metabolic stagnation in the placebo arm—constitute a clinically meaningful signal of hepatocellular stress mitigation and robust hepatic safety (Figure 4B).

### 3.5 Cortisol

Serum cortisol concentrations remained highly stable and strictly within the established morning reference range (6.4–22.8 µg/dL) throughout the 30-day intervention for both cohorts. In the EGT group, cortisol levels were 11.37 ± 2.85 µg/dL at Baseline and 11.81 ± 3.01 µg/dL at Day 30 (*p* = 0.64). The placebo group exhibited similar stability (Baseline: 14.07 ± 3.32 µg/dL; Day 30: 14.21 ± 3.26 µg/dL; *p* = 0.87). No significant between-group differences were detected at any timepoint, indicating that 30-day EGT supplementation does not disrupt the physiological normalization of the hypothalamic-pituitary-adrenal (HPA) axis in postpartum women (Figure 4B).

### 3.6 Correlations Between Change Scores

To comprehensively assess the relationships between subjective functional recovery and objective physiological markers, Pearson’s correlation coefficients were computed across the combined cohort utilizing change scores (Δ = Day 30 minus Baseline).

The ΔSF-36 total score was strongly and negatively correlated with ΔPSQI (r = −0.54, *p* < 0.05), ΔFS-14 (r = −0.64, *p* < 0.01), and the ΔTCM asthenia score (r = −0.64, *p* < 0.01). This tightly coupled cluster mathematically validates that the enhancement in broad quality of life is inherently driven by the specific resolution of sleep disturbances and profound systemic fatigue.

Furthermore, an exploratory analysis of biochemical change scores revealed that ΔCortisol is negatively correlated (r = −0.51, *p* < 0.05) with ΔALT, alongside a similar negative correlation with ΔAST (r = −0.47, *p* < 0.05). However, as serum cortisol concentrations remained stable throughout the intervention in both groups, this correlation should be interpreted with caution and may reflect statistical artifact rather than true physiological coupling, warranting further investigation in larger cohorts (Figure 5).

**Figure 5.**
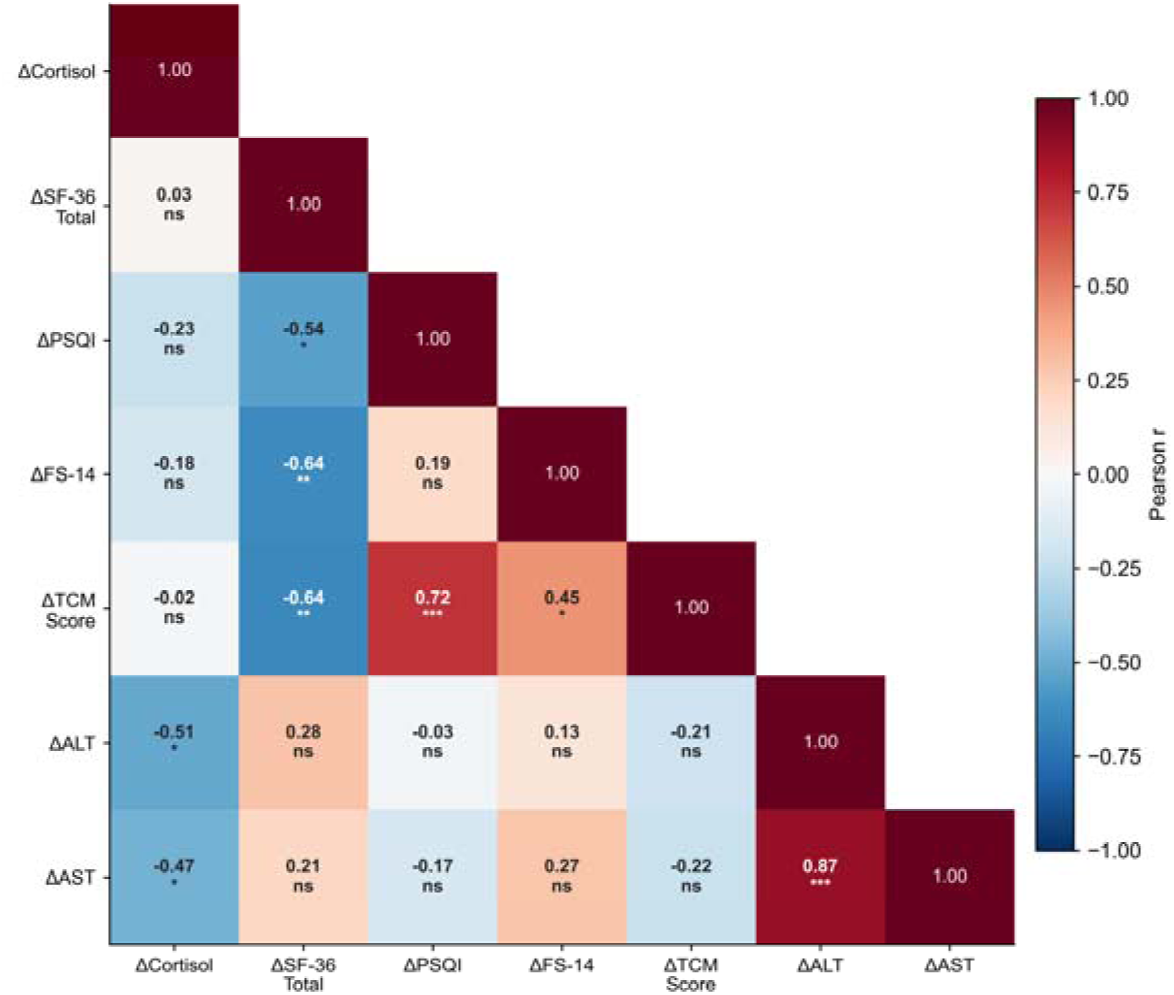
Correlation heatmap of clinical and biochemical change scores (Δ). Color intensity represents Pearson’s correlation coefficients (r). Significant correlations (p < 0.05) are highlighted, demonstrating the coupling between quality of life improvements and hepatic/sleep/fatigue recovery.

### 3.7 Safety

During the 30-day trial, daily EGT supplementation at 120 mg/day was exceptionally well-tolerated. Comprehensive hematological parameters—encompassing complete blood counts and core renal function indices (blood urea nitrogen and serum creatinine)—remained stable and strictly within normal reference ranges at all evaluation points.

A comprehensive summary of Treatment-Emergent Adverse Events (TEAEs) categorized by System Organ Class is presented in Table 3. Crucially, no participants reported instances of gastrointestinal intolerance, allergic reactions, or any other treatment-emergent symptoms. One case of unintended pregnancy was reported in the EGT group. However, following rigorous clinical evaluation, the trial physicians definitively determined this to be an unexpected medical event unrelated to the trial intervention. Comprehensive hematological and biochemical data demonstrating the excellent safety profile of EGT are presented in Supplementary Table S1.

**Table 3.**
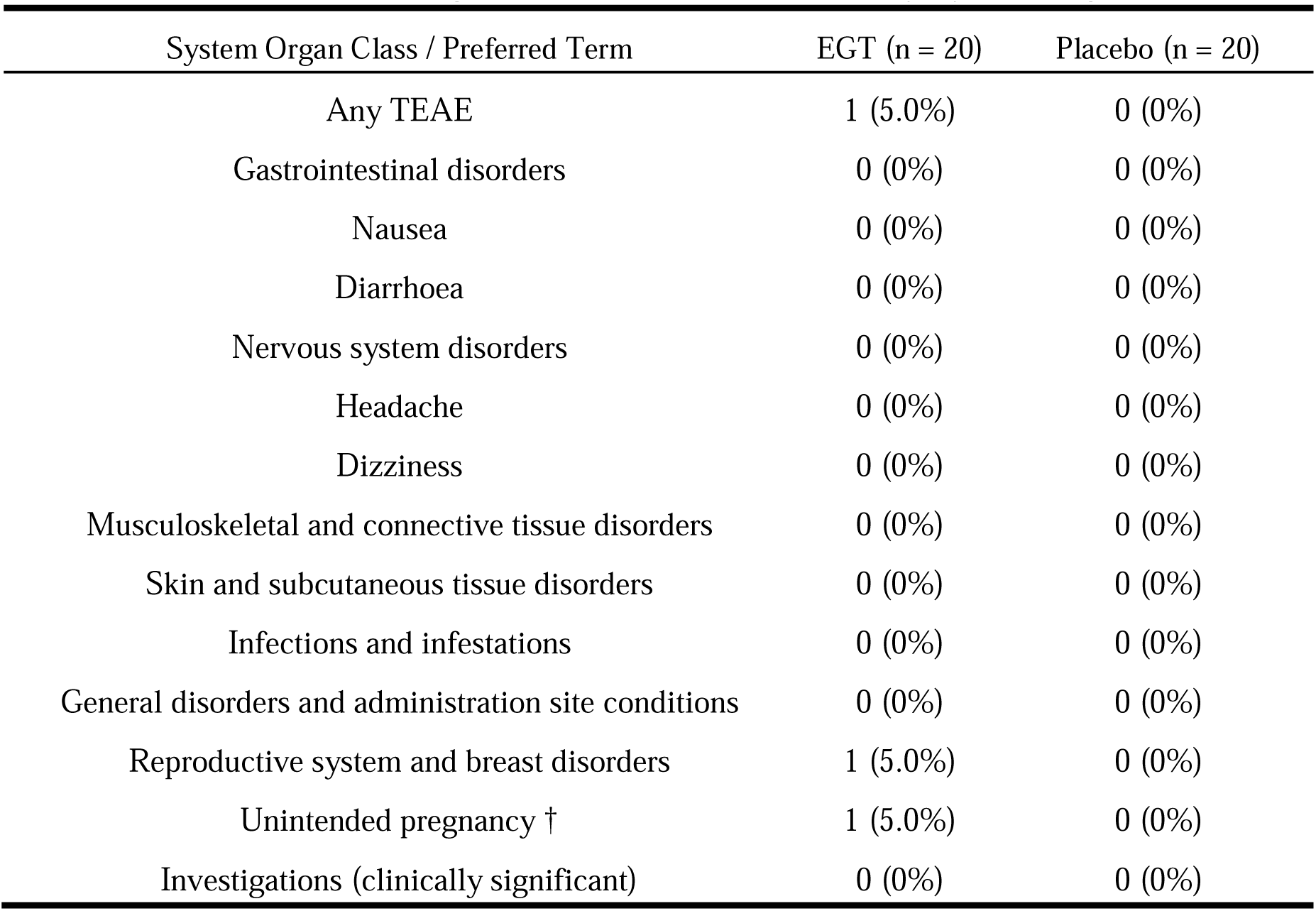
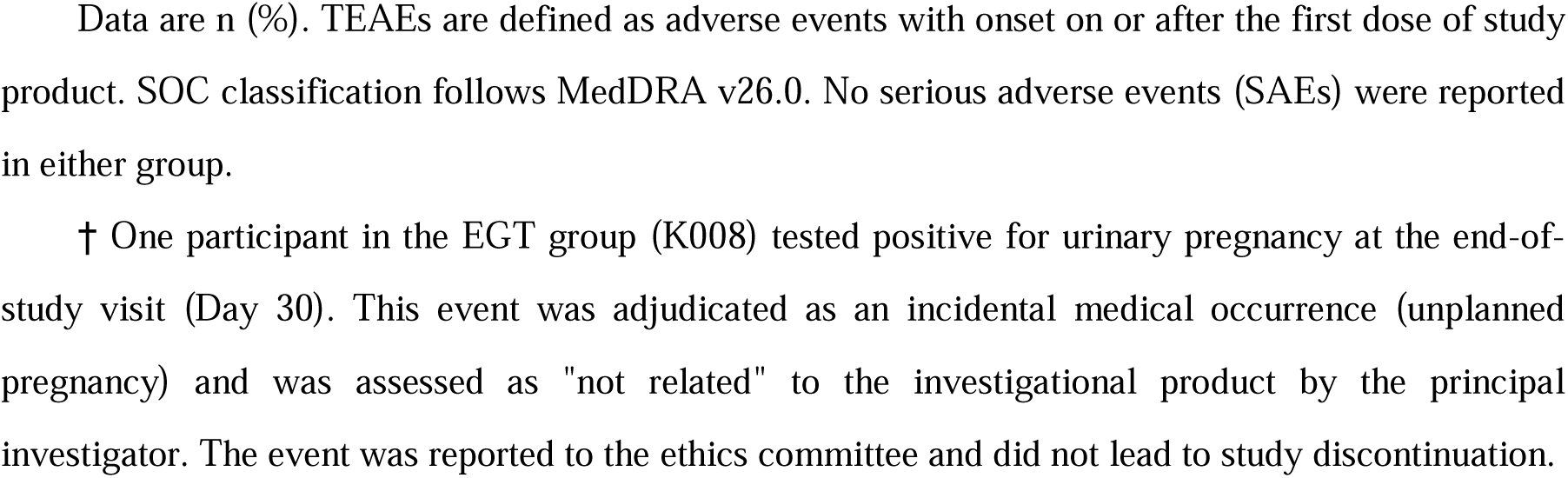
Treatment-Emergent Adverse Events (TEAEs) by System Organ Class.

| System Organ Class / Preferred Term | EGT (n = 20) | Placebo (n = 20) |
| --- | --- | --- |
| Any TEAE | 1 (5.0%) | 0 (0%) |
| Gastrointestinal disorders | 0 (0%) | 0 (0%) |
| Nausea | 0 (0%) | 0 (0%) |
| Diarrhoea | 0 (0%) | 0 (0%) |
| Nervous system disorders | 0 (0%) | 0 (0%) |
| Headache | 0 (0%) | 0 (0%) |
| Dizziness | 0 (0%) | 0 (0%) |
| Musculoskeletal and connective tissue disorders | 0 (0%) | 0 (0%) |
| Skin and subcutaneous tissue disorders | 0 (0%) | 0 (0%) |
| Infections and infestations | 0 (0%) | 0 (0%) |
| General disorders and administration site conditions | 0 (0%) | 0 (0%) |
| Reproductive system and breast disorders | 1 (5.0%) | 0 (0%) |
| Unintended pregnancy † | 1 (5.0%) | 0 (0%) |
| Investigations (clinically significant) | 0 (0%) | 0 (0%) |

Data are n (%). TEAEs are defined as adverse events with onset on or after the first dose of study product. SOC classification follows MedDRA v26.0. No serious adverse events (SAEs) were reported in either group.
† One participant in the EGT group (K008) tested positive for urinary pregnancy at the end-of-study visit (Day 30). This event was adjudicated as an incidental medical occurrence (unplanned pregnancy) and was assessed as "not related" to the investigational product by the principal investigator. The event was reported to the ethics committee and did not lead to study discontinuation.

## 4. Discussion

This randomized, double-blind, placebo-controlled pilot trial evaluated the efficacy of a 30-day EGT supplementation (120 mg/day) in postpartum women experiencing reduced quality of life. Both the EGT and placebo cohorts demonstrated statistically significant within-group improvements across all primary and secondary subjective outcome measures. Although absolute between-group differences at Day 30 did not reach formal statistical significance in unadjusted analyses, the robust improvements in the placebo arm are consistent with the well-documented placebo effect and the natural physiological recovery trajectory of the postpartum period [1, 15]. The primary contribution of this study lies in characterizing the selective therapeutic signals of EGT, notably the accelerated onset of functional recovery and the concurrent optimization of hepatic safety markers.

The most compelling and clinically profound finding of this trial is the temporal acceleration of functional recovery in the EGT group. Participants receiving EGT achieved statistically significant within-group improvements in sleep architecture (PSQI) and systemic fatigue (FS-14) as early as Day 15, a critical timepoint where the placebo cohort exhibited no significant changes. Most notably, sub-scale analysis revealed a significant between-group superiority for EGT in alleviating mental fatigue at Day 15 (nominal *p* = 0.0313). This rapid, two-week acceleration in cognitive and physical restoration is highly relevant to postpartum care, as it may actively reduce the downstream risk of postpartum mood disorders and substantially improve maternal caregiving capacity [1, 4]. While the precise neurobiological mechanisms require further elucidation, EGT’s targeted antioxidant actions within the central nervous system may effectively mitigate neuroinflammation-driven cognitive fatigue. Furthermore, advanced statistical modeling (ANCOVA) confirmed that, even after adjusting for baseline disparities, EGT consistently maintained a positive numerical superiority in overall quality of life (SF-36) at Day 30.

As a critical secondary finding supporting this functional recovery, EGT demonstrated a selective reduction in hepatic transaminases. ALT levels declined by 30.42% (*p* = 0.019) and AST by 17.44% (*p* = 0.006) in the EGT arm, while neither parameter reached significance in the placebo group. The postpartum liver is subjected to a heightened metabolic load due to the rapid clearance of gestational steroid hormones and sustained mitochondrial ROS production [6]. EGT’s unique biological property—specifically its capacity to concentrate in mitochondria via the OCTN1 transporter and preserve electron transport chain integrity—provides a robust mechanistic rationale for the observed transaminase reductions [7, 9]. We hypothesize that this hepatic optimization is intrinsically linked to the alleviation of systemic fatigue; by mitigating hepatocellular stress, EGT may enhance overall mitochondrial bioenergetics, thereby supplying the necessary energetic foundation for rapid physical and mental restoration (a “hepatic-energetic” axis).

The parallel improvement observed in the Traditional Chinese Medicine (TCM) Qi-Blood deficiency scores further highlights the utility of a multi-framework approach to assessing postpartum recovery. The robust reduction in systemic asthenia within the EGT group (−42.78% vs. −31.03% in placebo) aligns conceptually with the TCM principle of “tonifying Qi.” In modern biological terms, this conceptual framework correlates strongly with Western metrics of reduced fatigue and overall metabolic efficiency [17]. This convergence of Eastern symptomatology and Western clinical metrics represents a methodological strength of this study, facilitating a more holistic and culturally sensitive understanding of postpartum functional restoration.

Serum cortisol concentrations remained stable and within normal physiological ranges across both cohorts, indicating that 30-day EGT supplementation does not substantially modify HPA axis activity in this population. This is biologically plausible, as the postpartum HPA axis undergoes a protracted normalization process over several months; a 30-day intervention is likely insufficient to alter the neuroendocrine set-point, particularly when baseline levels are already within normal limits [16]. The cross-domain correlations between change scores (Δ) provide compelling evidence for the construct validity of our outcome battery. Strong negative correlations between ΔSF-36 and both ΔPSQI (r = −0.54) and ΔFS-14 (r = −0.64) confirm that enhanced quality of life is tightly coupled to objective reductions in sleep disturbance and systemic fatigue.

Regarding safety, the excellent tolerability and improved hepatic profile observed in this postpartum cohort is consistent with EGT’s established safety profile in healthy adults [11]. This clinical safety is further underscored by its regulatory status as Generally Recognized as Safe (GRAS) by the US FDA and its approval as a novel food ingredient by the European Food Safety Authority (EFSA).

This study has several limitations. First, the sample size (n = 20 per arm) represents an exploratory pilot design, lacking the power to detect definitive between-group differences in subjective metrics without covariate adjustment. Future trials would require approximately 80–130 participants per arm to achieve 80% power based on the observed effect sizes (Cohen’s d ≈ 0.30–0.50). Second, the single-center, 30-day follow-up may be insufficient to capture the full trajectory of hepatic and neuroendocrine normalization. Additionally, a numerically meaningful difference in baseline morning serum cortisol was observed between groups (EGT: 11.37 ± 2.85 vs. Placebo: 14.07 ± 3.32 µg/dL; p = 0.08), which, while not reaching statistical significance, may reflect residual baseline imbalance in neuroendocrine stress reactivity. Although cortisol remained stable within normal limits in both arms throughout the intervention—confirming EGT’s safety with respect to HPA axis function—this baseline disparity limits the interpretability of inter-group cortisol comparisons. Future confirmatory trials should employ pre-stratified randomization on cortisol levels and ANCOVA-adjusted neuroendocrine analyses to more rigorously characterize EGT’s effects on HPA axis normalization. Finally, the absence of direct oxidative stress biomarkers (e.g., MDA, SOD) and plasma EGT quantification restricts deeper mechanistic interpretations.

## 5. Conclusion

This randomized, double-blind, placebo-controlled pilot trial demonstrates that EGT supplementation (120 mg/day) for 30 days is safe and exceptionally well-tolerated in postpartum women. Significant within-group improvements were observed across all primary and secondary subjective recovery endpoints, including quality of life (SF-36, +35.94%, *p* = 0.0006), sleep quality (PSQI, −29.24%, *p* = 0.0001), fatigue (FS-14, −27.78%, p = 0.0011), and systemic asthenia (−42.78%, *p* = 0.0012). Crucially, EGT demonstrated an accelerated onset of action, yielding significant between-group superiority in relieving mental fatigue by Day 15. The selective reduction in hepatic transaminases (ALT −30.42%; AST −17.44%) acts as an additional physiological benefit, further confirming its excellent safety profile. While absolute between-group differences at study completion did not reach significance—consistent with the trial’s exploratory design—these robust and early-onset clinical signals warrant confirmation in adequately powered, multicenter clinical trials.

## Data Availability

The datasets generated and analyzed in this study are available from the corresponding author on reasonable request.

## Author Contributions

C.G., J.C., and F.S. coordinated clinical operations and project administration. W.D. supervised the overall study. S.Z. supervised clinical assessments at the study site. W.L., C.G., J.C., G.X., and T.T. performed data and statistical analyses. W.L. drafted the manuscript. All authors reviewed and approved the final manuscript.

