## Supplementary_Materials for "Efficacy of Gene III^®^ L-Ergothioneine Capsules on Postpartum Fatigue, Sleep Quality, and Quality of Life: A Randomized, Double-Blind, Placebo-Controlled Trial"

**Supplementary Material**

Supplementary Figure S1

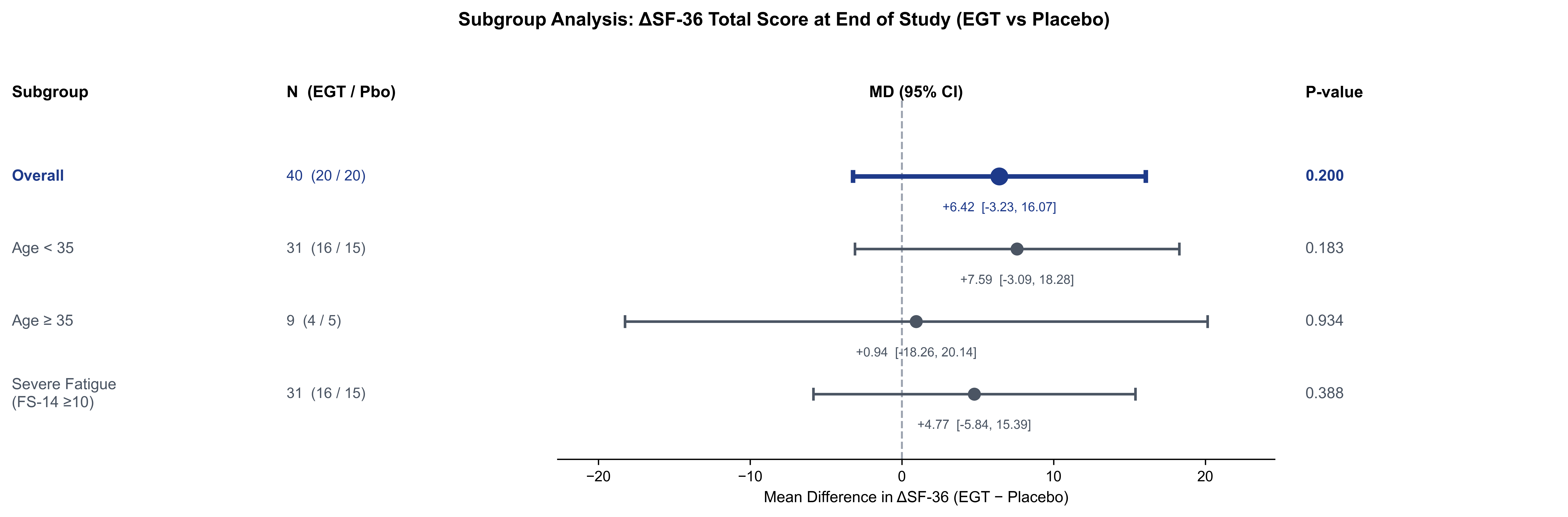
Forest plot of SF-36 total score change (Δ, Day 30 − Baseline) across pre-specified subgroups. Point estimates represent between-group mean differences (EGT minus Placebo) with 95% confidence intervals. Subgroups: Overall (n = 40); Age < 35 years (n = 31); Age ≥ 35 years (n= 9); Severe fatigue at baseline (FS-14 ≥ 10, n = 31).

Supplementary Table S1

Hematological and Biochemical Safety Parameters at Baseline and Day 30

| **Parameters** | **EGT Group (n = 20)** | | **Placebo Group (n = 20)** | | **Between-group P-value** |
| --- | --- | --- | --- | --- | --- |
| **Parameter (unit)** | **Baseline** | **Day 30** | **Baseline** | **Day 30** | **P-value (Day 30)** |
| WBC, ×10⁹/L | 6.20 ± 1.29 | 6.10 ± 1.71 | 6.22 ± 2.00 | 6.13 ± 1.54 | 0.957 |
| RBC, ×10¹²/L | 4.77 ± 0.51 | 4.69 ± 0.53 | 4.65 ± 0.33 | 4.61 ± 0.38 | 0.594 |
| Hb, g/L | 135.2 ± 10.5 | 133.2 ± 12.0 | 134.4 ± 9.4 | 134.1 ± 12.2 | 0.815 |
| PLT, ×10⁹/L | 239.4 ± 74.2 | 258.9 ± 67.1 | 249.6 ± 66.3 | 253.8 ± 52.8 | 0.795 |
| ALT, U/L | 17.8 ± 16.6 | 12.3 ± 6.6 | 20.9 ± 19.7 | 22.7 ± 30.9 | 0.151 |
| AST, U/L | 19.5 ± 9.3 | 16.1 ± 4.2 | 19.4 ± 6.8 | 18.7 ± 9.9 | 0.288 |
| TBIL, μmol/L | 12.25 ± 4.10 | 11.05 ± 3.78 | 10.49 ± 3.90 | 10.67 ± 3.97 | 0.755 |
| Scr, μmol/L | 49.8 ± 6.5 | 50.1 ± 6.1 | 49.7 ± 7.0 | 51.6 ± 8.3 | 0.527 |
| BUN, mmol/L | 4.55 ± 0.95 | 4.33 ± 1.03 | 4.72 ± 1.47 | 4.55 ± 1.23* | 0.552 |

Data are presented as Mean ± SD. Between-group comparisons at Day 30 were performed using independent-samples t-test. All P-values > 0.05, indicating no statistically significant differences between groups in any hematological or biochemical parameter at Day 30, confirming the safety of EGT supplementation.
One BUN value (participant K029, Day 30, EGT group: 88 mmol/L) was identified as a probable data-entry error (clinical reference range: 3.6–7.1 mmol/L) and was excluded from the BUN analysis (effective n = 19 for EGT group BUN at Day 30).
Abbreviations: WBC, white blood cell count; RBC, red blood cell count; Hb, hemoglobin; PLT, platelet count; ALT, alanine aminotransferase; AST, aspartate aminotransferase; TBIL, total bilirubin; Scr, serum creatinine; BUN, blood urea nitrogen; EGT, ergothioneine; SD, standard deviation.
